# Improving bystander automated external defibrillation application in Singapore: An 11-year population-based living-laboratory study

**DOI:** 10.64898/2026.05.20.26353744

**Authors:** John T. Bokman, Shaynna Ee, Stephanie Fook-Chong, Nur Shahidah, Benjamin SH Leong, Michael YC Chia, Yohei Okada, Marcus EH Ong, Fahad Javaid Siddiqui, the Singapore PAROS Investigators

## Abstract

**Background:** Bystander automated external defibrillator (BAED) use improves out-of-hospital cardiac arrest (OHCA) outcomes but remains uncommon globally. This study evaluated the outcomes of Singapore’s 11-year public-access AED expansion and volunteer-responder implementation in terms of trends in BAED application, associated factors, and clinical outcomes.

**Methods:** This population-based, retrospective cohort study used Singapore Pan-Asian Resuscitation Outcomes Study (SG-PAROS) data (2010–2020) for adult, non-traumatic OHCAs. The primary outcome was bystander AED application. Multivariable logistic regression identified factors associated with use. Secondary outcomes included favorable neurological status (CPC 1–2), survival to discharge, and prehospital return of spontaneous circulation (ROSC).

**Results:** Of 21,439 included OHCA cases (median age 70.0 years; 63.8% male), BAED application increased from 1.7% to 9.6% over 11 years, with a corresponding increase in overall survival from 2.4 to 4.0%. Malay ethnicity (aOR 1.25, 1.06–1.49), calendar year (aOR 1.26, 1.22–1.29), and delayed emergency medical services (aOR 1.24, 1.06–1.45) were positive predictors of BAED application. Conversely, BAED application was lower among females (aOR 0.80, 95% CI 0.69–0.94), at night (aOR 0.69, 0.56–0.86), and in residential settings (aOR 0.06, 0.05–0.07).

Volunteer arrival strongly increased application (aOR 4.16, 3.41–5.09), with a significant interaction (p<0.001); the effect was greater in residential (aOR 7.38, 5.81–9.38) than non-residential settings (aOR 1.71, 1.22–2.40). AED use predicted favorable neurological outcome (aOR 2.80, 2.24–3.50; NNT 8.7), survival (aOR 2.30, 1.89–2.80), and ROSC (aOR 2.11, 1.81–2.46).

**Conclusion:** Over 11 years, we saw a significant increase in BAED application and favorable neurological survival. This success was associated with the implementation of an integrated strategy combining widespread AED deployment, national training, and smartphone-activated volunteer responders. Singapore’s experience provides a scalable model for urban centers seeking to expand their AED strategy.

## Introduction

Out-of-hospital cardiac arrest (OHCA) is a major global public health concern with an overall survival rate less than 10%.^1^ Early defibrillation using an automated external defibrillator (AED) by bystanders substantially improves survival and neurological outcomes.^2–6^ Despite its importance, bystander AED (BAED) use remains low worldwide, with reported rates generally ranging from 2% to 20% depending on region and system characteristics.^7^

Over the past two decades, several regions have demonstrated that coordinated public-access defibrillation strategies can substantially improve BAED use and OHCA outcomes. In Denmark, nationwide initiatives combining large-scale AED deployment with linkage to emergency dispatch systems and volunteer responder activation have led to marked increases in bystander defibrillation and survival.^8^ In the United States, large registry studies have shown that BAED use in public settings is associated with significantly improved survival and neurological outcomes.^2^ In Sweden, smartphone-based activation of trained volunteer responders has further strengthened early response by facilitating rapid initiation of cardiopulmonary resuscitation (CPR) and retrieval of nearby AEDs prior to emergency medical services arrival.^9^ These examples highlight how integrated approaches that combine data-driven AED deployment, real-time registry integration, and community responder mobilization can enhance early defibrillation and improve survival outcomes.

Singapore has experienced a substantial rise in OHCA incidence, increasing from 25 to 74 per 100,000 population between 2010 and 2021.^10^ During this period, Singapore has sought to build a living laboratory for OHCA by implementing a series of interventions to increase bystander CPR (BCPR) and BAED use. The impact of these programs on BCPR has previously been described.^11^ These include widespread CPR/AED training, dense deployment of public-access AEDs (including residential estates), and development of a national AED registry.^12–14^ In addition, Singapore has implemented the myResponder mobile application, which crowdsources nearby volunteer community first responders and provides real-time AED location guidance.^12^ Early evidence suggests that myResponder activations are associated with increased bystander CPR and facilitates AED retrieval.^15,16^ These efforts to further develop AED strategies, both globally and in Singapore, reflect a broader shift toward integrated, community-based OHCA response systems.

To determine next steps for improving BAED use in Singapore, it is essential to reflect on the effectiveness of current interventions and identify opportunities for growth. One approach to investigate BAED use is to explore the factors associated with application. Prior studies have established the positive association of volunteer alert mobile applications and public-access AED programs on BAED and OHCA outcomes.^4,17,18^ Several barriers to BAED application have also been identified in other settings, including female sex, certain ethnic minorities, nighttime arrests, and weekend arrests.^19–21^ Notably, BAED application has been found to be significantly less likely in residential locations across numerous regions.^21–23^

It remains unclear whether these facilitators and barriers observed elsewhere apply in Singapore, or if local characteristics, such as the country’s unique racial composition or the *myResponder* system, shape BAED use in distinct ways.^24^ Clarifying how demographic, temporal, and location-based factors influence BAED application will be helpful to refine AED strategies and identify targeted opportunities to improve early defibrillation.

The objective of this study was to characterize patterns of BAED application in Singapore, usage trends over time, identify factors associated with BAED application, and evaluate the relationship between BAED application and clinical outcomes.

## Methods

### Ethics

The study protocol was reviewed and exempted from individual informed consent by the Centralized Institutional Review Board and Domain Specific Review Board in Singapore for the SG-PAROS database (ref no: 2010/270/C, 2013/604/C, 2013/00929 and 2018/2937). All patients were de-identified prior to analysis.

### Study Design and Setting

This observational study analyzed data from the Singapore cohort of the Pan-Asian Resuscitation Outcomes Study (PAROS). PAROS is a population-based registry of out-of-hospital cardiac arrests system that integrates data from emergency medical services (EMS), dispatch, and receiving hospitals using a standardized Utstein template.^25–27^ Data entry is performed by trained EMS personnel and verified through automated logical checks within the registry database; incomplete or inconsistent entries undergo follow-up clarification before finalization. Available variables include demographic characteristics, arrest circumstances, bystander interventions, EMS response times, and survival and neurologic outcomes. Data from April 1, 2010, to December 31, 2020, were included.

In Singapore, emergency medical services are provided by the Singapore Civil Defence Force, which operates a nationwide ambulance-based response system. Over the study period, multiple coordinated public health initiatives were implemented to strengthen early community response to OHCA and increase bystander AED use. These included large-scale deployment of public-access AEDs in which defibrillators were placed in high-traffic locations and in the ground-floor lobby of every alternate public housing block.^14^ In parallel, national CPR and AED training campaigns were expanded across schools, workplaces, and community organizations to increase layperson readiness and normalize bystander intervention.^12^ In addition, the myResponder mobile application was introduced in 2015 as a smartphone-activated volunteer responder system that alerts nearby trained individuals to suspected cardiac arrests and provides real-time geolocation of both the patient and the nearest registered AED.^15,16^ The application will alert pedestrian volunteers within 400 meters of an arrest and responders in a vehicle within 1 kilometer.

### Study Participants and Definitions

Adult (age ≥ 18) patients with OHCA of non-traumatic etiology were eligible for inclusion. Cases were excluded if the arrest was witnessed by EMS personnel or occurred in the ambulance as these scenarios lacked an opportunity for bystander AED use. Patients were also excluded if key data were missing, including bystander AED status, neurologic outcome, or required covariates for multivariable analysis.

A bystander was defined according to the Utstein definition as a person who is present at the scene or alerted to the arrest but not dispatched as part of an organized emergency response system.^27^ Bystanders include off-duty healthcare professionals and volunteer community responders. Bystander AED application was defined as any instance in which an AED was placed on the patient by a bystander, regardless of whether a shock was delivered or whether the AED pads were placed correctly. EMS response time was defined as the interval in minutes from call received to EMS arrival at the patient’s side with delayed EMS response being greater than or equal to nine minutes based on industry standard.^28^

### Data Collection

From the PAROS registry, the following variables were extracted: age, gender, ethnicity, date and time of arrest, arrest location (residential vs public), witness status, bystander CPR, EMS response time (interval in minutes from call to EMS arrived at patient side), application of a bystander AED, initial rhythm, and outcome measures including survival to hospital discharge or 30 days, prehospital return of spontaneous circulation (ROSC), and Cerebral Performance Category (CPC).^27,29^

Data from the Singapore Civil Defence Force myResponder system database (January 1, 2016 to December 31, 2020) were matched to cases in PAROS to capture volunteer responder activity, specifically whether an alert was sent, accepted, and whether a responder arrived on scene. For all cases prior to 2016, these myResponder variables were coded as zero to reflect the absence of the system.

### Outcome Measures

The primary outcome was bystander AED use, defined as the application of AED pads to the patient prior to EMS arrival regardless of whether a shock was delivered. Secondary outcomes were favorable neurological outcome, defined as a CPC score of 1–2 at discharge or 30 days post-arrest, survival to hospital discharge or 30 days post-arrest, and prehospital ROSC, defined as regaining of a palpable pulse.

### Statistical Analysis

Descriptive statistics were calculated using medians and interquartile ranges (IQRs) for continuous variables and counts with percentages for categorical variables. Univariate logistic regression analyses were first conducted for each covariate. Variables with a p-value < 0.2, as well as those considered clinically or theoretically important, were included in the multivariable model. Multivariable logistic regression was performed to identify factors associated with bystander AED use. Covariates in the model were: gender, age, ethnicity, time of day, location of arrest, witness status, myResponder volunteer arrived, EMS response time, and year.

Multivariable logistic regression models assessed the association between BAED and clinical outcomes, including favorable neurologic status (CPC 1–2), overall survival, and prehospital ROSC. These models adjusted for age, gender, ethnicity, weekend, time of day, arrest location, witness status, EMS response time, bystander CPR, and study year.

An observationally derived Number Needed to Treat (NNT) was estimated from adjusted outcome probabilities to quantify the association between bystander AED application and favorable neurological survival, with 95% confidence intervals.

To explore potential effect modification, we tested for two-way interactions within our primary models. For the AED application model, we assessed the interaction between arrest location and potential modifiers: patient age category and myResponder volunteer arrival. For the survival model, we evaluated for interactions between bystander AED application and four potential modifiers: arrest location, time of arrest, witness status, and volunteer arrival. Interaction terms were assessed using the likelihood ratio test, with statistical significance set at p < 0.05. Where significant interaction was identified, we performed stratified analyses to report setting-specific adjusted odds ratios (aOR).

All analyses were performed using Python (version 3.11.5) with pandas (v2.0.3), NumPy (v1.24.3), and statsmodels (v0.14.0). All tests were two-tailed with statistical significance set at P < 0.05.

## Results

### Cohort Characteristics

A total of 21,439 adult (age ≥ 18) OHCA cases met inclusion criteria, of which 1,325 (6.2%) received bystander AED application prior to EMS arrival (Figure 1). Compared with patients who did not receive AED application, those who did were younger (median age 64.0 years [IQR 54.0–77.0] vs 71.0 [59.0–82.0]) and more likely to be male (78.5% vs 62.8%) compared with those who did not receive AED application (Table 1).

**Table 1.** Case characteristics. Baseline demographic, arrest, and prehospital characteristics of adult (age ≥ 18) non-traumatic out-of-hospital cardiac arrest cases in Singapore from 2010 to 2020, stratified by whether a bystander applied an automated external defibrillator (AED) prior to EMS arrival.

| Characteristics | BAED applied<br>(n = 1,325) | BAED not applied (n=20,114) | Overall<br>(n =21,439) |
| --- | --- | --- | --- |
| Age (years)* | 64.0 [54.0–76.0] | 71.0 [59.0–82.0] | 70.0 [58.0–81.0] |
| Age (years)† |  |  |  |
| <55 | 350 (26.4%) | 3617 (18.0%) | 3967 (18.5%) |
| 55-64 | 325 (24.5%) | 3815 (19.0%) | 4140 (19.3%) |
| 65-74 | 274 (20.7%) | 4366 (21.7%) | 4640 (21.6%) |
| 75-84 | 197 (14.9%) | 4684 (23.3%) | 4881 (22.8%) |
| $\geq 85$ | 179 (13.5%) | 3632 (18.1%) | 3811 (17.8%) |
| Sex, male | 1040 (78.5%) | 12639 (62.8%) | 13679 (63.8%) |
| Ethnicity |  |  |  |
| Chinese | 836 (63.1%) | 13959 (69.4%) | 14795 (69.0%) |
| Malay | 221 (16.7%) | 3110 (15.5%) | 3331 (15.5%) |
| Indian | 182 (13.7%) | 2145 (10.7%) | 2327 (10.9%) |
| Other | 86 (6.5%) | 900 (4.5%) | 986 (4.6%) |
| Weekend | 362 (27.3%) | 5935 (29.5%) | 6297 (29.4%) |
| Time of day |  |  |  |
| Day (06:00-17:59) | 890 (67.2%) | 11738 (58.4%) | 12628 (58.9%) |
| Evening (18:00-23:59) | 324 (24.5%) | 5348 (26.6%) | 5672 (26.5%) |
| Night (00:00-05:59) | 111 (8.4%) | 3028 (15.1%) | 3139 (14.6%) |
| Location, residence | 312 (23.5%) | 16052 (79.8%) | 16364 (76.3%) |
| Witnessed arrest | 788 (59.5%) | 10420 (51.8%) | 11208 (52.3%) |
| Initial rhythm category |  |  |  |
| Non-shockable | 810 (61.1%) | 16844 (83.7%) | 17654 (82.3%) |
| Shockable | 508 (38.3%) | 3152 (15.7%) | 3660 (17.1%) |
| Unknown | 7 (0.5%) | 118 (0.6%) | 125 (0.6%) |
| Bystander defibrillation given | 434 (32.8%) | 11 (0.1%) | 445 (2.1%) |
| myResponders arrived | 196 (14.8%) | 1202 (6.0%) | 1398 (6.5%) |
| EMS response delayed ($\geq 9$ minutes) | 1072 (80.9%) | 15487 (77.0%) | 16559 (77.2%) |
| Dispatch-assisted CPR | 420 (29.7%) | 8104 (36.0%) | 8524 (35.7%) |
| Year of arrest |  |  |  |
| 2010-2011 | 39 (2.9%) | 2075 (10.3%) | 2114 (9.9%) |
| 2012-2014 | 123 (9.3%) | 4388 (21.8%) | 4511 (21.0%) |
| 2015-2017 | 350 (26.4%) | 6169 (30.7%) | 6519 (30.4%) |
| 2018-2020 | 813 (61.4%) | 7482 (37.2%) | 8295 (38.7%) |

\*Continuous variables are presented as a median [interquartile range]
†Categorical variables as a number (percentage).

**Figure 1.**
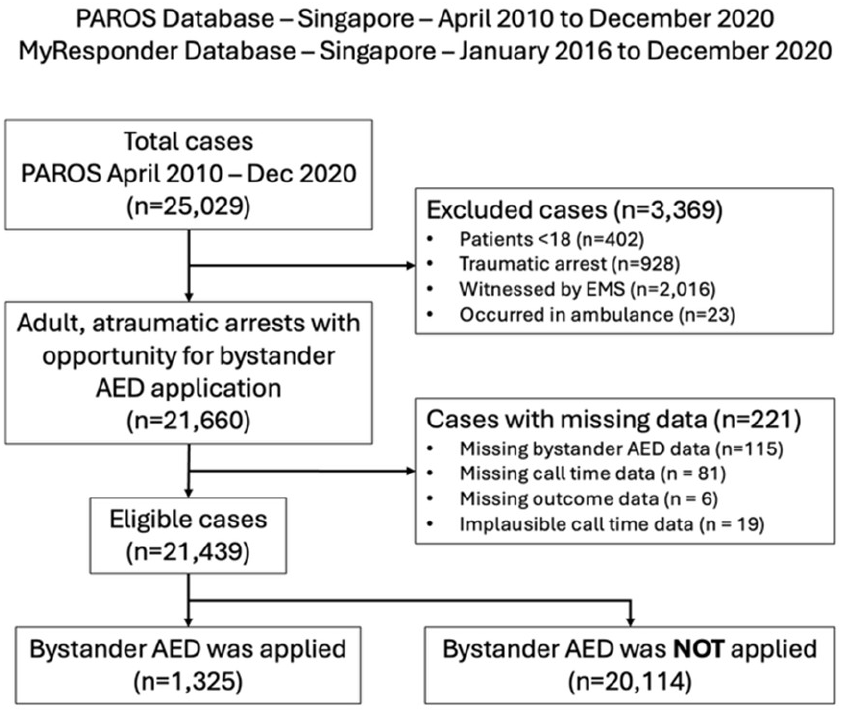
Flow diagram of study cohort selection illustrating inclusion and exclusion criteria. Applied to out-of-hospital cardiac arrest cases from the PAROS registry in Singapore between 2010 and 2020, resulting in the final analytic cohort.

Arrest location and timing were strongly associated with AED use. Only 23.5% of AED applications occurred in residential locations, compared with 79.8% of cases without AED use. AED application was also more common in daytime hours (67.2% vs 58.4%) compared with evening or nighttime. Shockable initial rhythms were substantially more frequent among cases with AED use (38.3% vs 15.7%). Bystander presence, defibrillation by lay rescuers, and myResponder volunteer arrival were all associated with higher AED application rates. Delayed EMS response (≥9 minutes) was slightly more common among cases with AED application (80.9% vs 77.0%), while dispatch-assisted CPR was less frequent (29.7% vs 36.0%).

### Trends

The proportion of OHCAs receiving bystander AED application increased over the study period (1.7% to 9.6%), with a parallel increase in bystander defibrillation (1.0% to 3.6%) (Figure 2). A corresponding increase in survival to discharge/30-day was also observed from 2.4% to 4.0% with a max of 5.4% prior to the COVID-19 pandemic. While BAED application rates in non-residential locations surged from 5.2% to 25.6%, residential application rates increased from 0.1% to 4.6%, resulting in an absolute gap that widened from 5.1% to 21.0% over the decade (Figure 3). This widening difference in bystander intervention was closely mirrored by a divergence in clinical outcomes. Favorable neurological survival in non-residential OHCAs nearly tripled, rising from 2.9% in 2010 to 8.6% in 2020. In contrast, neurological survival in residential settings increased marginally from 1.0% to 1.2% over the same timeframe.

**Figure 2.**
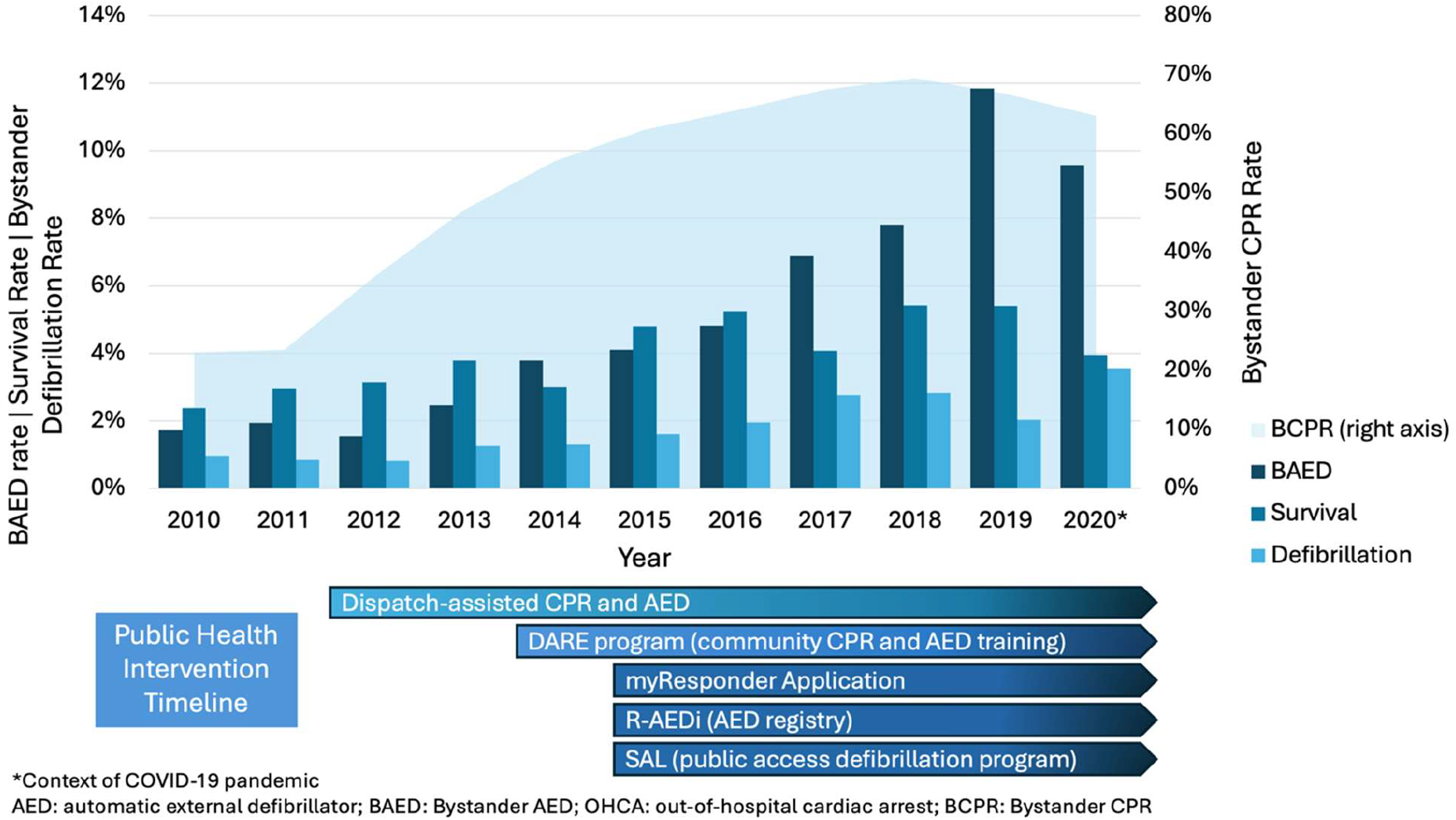
Bystander AED application, defibrillation, survival, and bystander CPR rate trends along public intervention timeline. Temporal trends in bystander interventions and patient survival of adult (age ≥ 18) out-of-hospital cardiac arrest cases in Singapore from 2010 to 2020. BAED application, bystander delivered defibrillation, and survival on the left axis. Bystander CPR on the right axis. Timeline of major public health interventions targeting BAED included for context.

**Figure 3.**
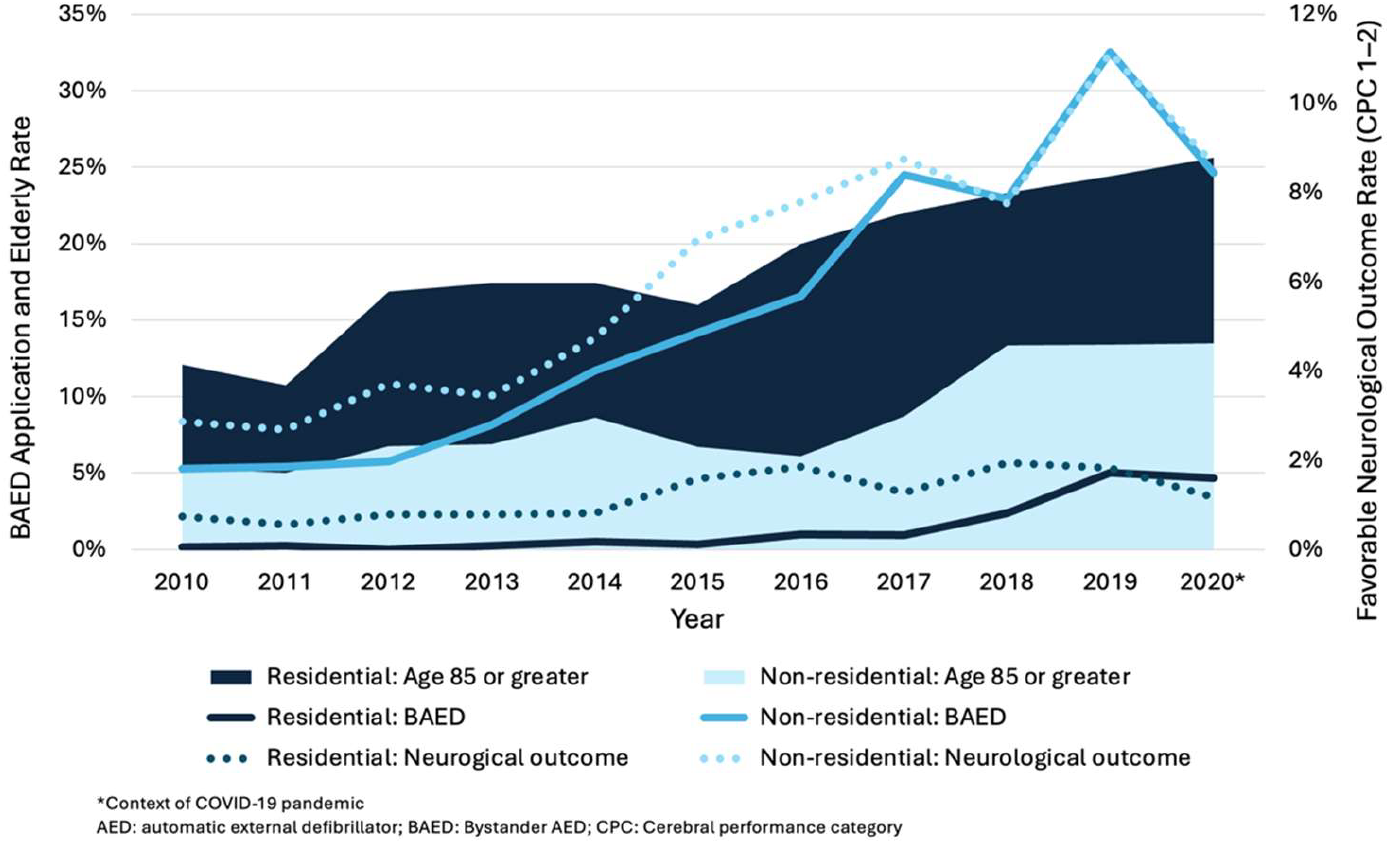
Trends in bystander AED application, favorable neurological outcome, and rate of elderly arrest by location (2010–2020). Trends are stratified by residential (dark blue) and non-residential (light blue) settings. Left axis: Bystander AED application and the proportion of patients greater than 85 years of age. Right axis: Favorable neurological outcomes (CPC 1–2).

### Factors Associated with Bystander AED Application

Multivariable logistic regression identified several independent determinants of bystander AED application (Figure 4). Female sex was associated with lower odds of AED use (adjusted odds ratio [aOR] 0.80, 95% confidence interval [CI] 0.69–0.94, p=0.005). Among ethnic groups, Malay individuals had slightly higher odds compared with Chinese patients (aOR 1.25, 95% CI 1.06–1.49, p=0.010), while other groups were not significantly associated with AED use.

**Figure 4.**
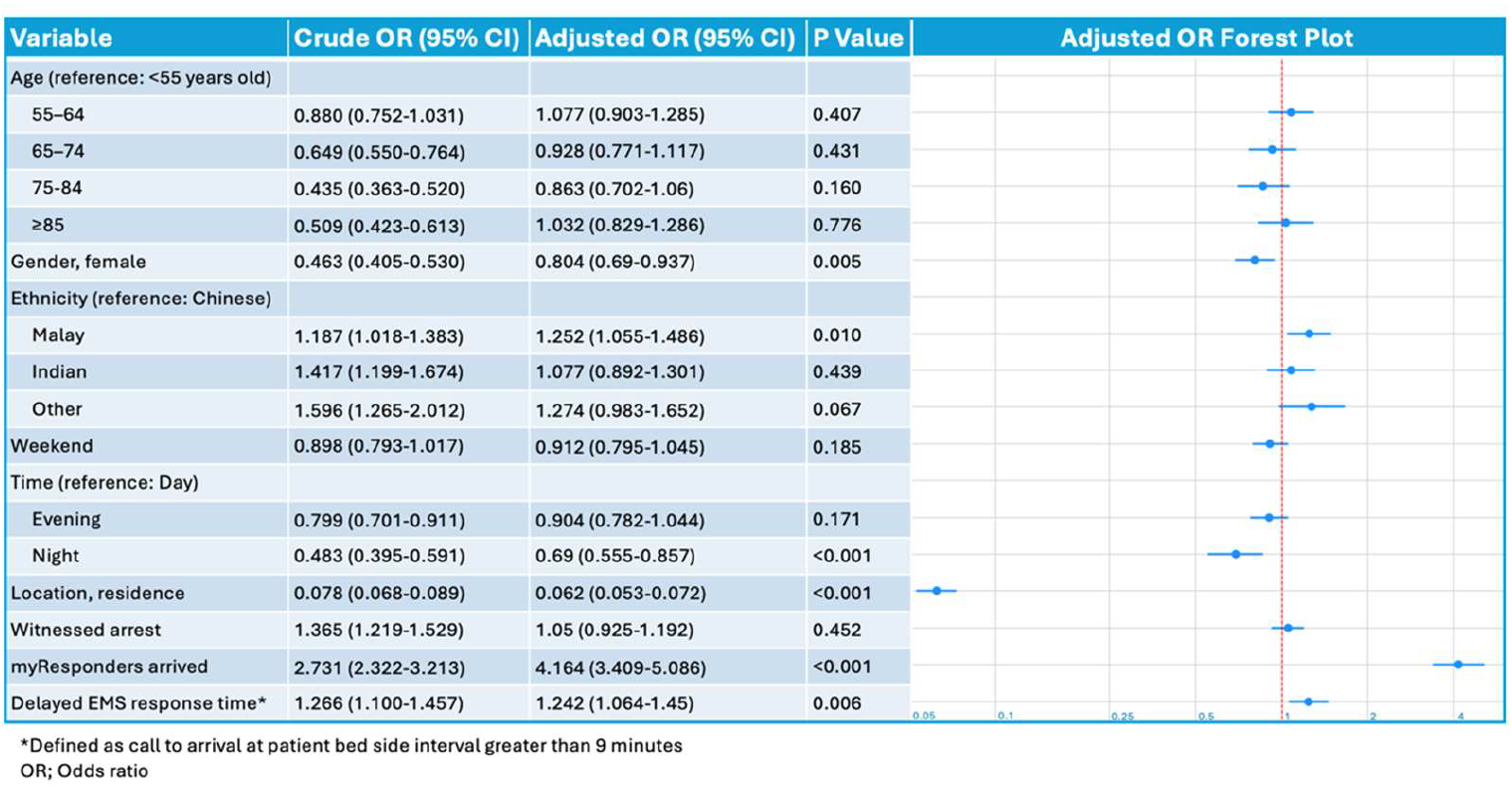
Univariate and multivariable analysis of factors associated with bystander automatic external defibrillator application. Crude and adjusted odds ratios from logistic regression analyses evaluating demographic, temporal, situational, and system-level factors associated with bystander AED application prior to EMS arrival among adult (age ≥ 18) out-of-hospital cardiac arrest cases.

Contextual factors strongly influenced AED use. AED application was markedly less likely in residential settings (aOR 0.06, 95% CI 0.05–0.07, p<0.001) and during nighttime hours (aOR 0.69, 95% CI 0.56–0.86, p<0.001). Later calendar year was independently associated with higher AED use (aOR 1.26 per year, 95% CI 1.22–1.29, p<0.001).

Delayed EMS response (≥9 minutes) was associated with a modestly higher likelihood of bystander AED application (aOR 1.24, 95% CI 1.06–1.45, p=0.006). Arrival of a smartphone-activated volunteer responder was strongly associated with BAED application (aOR 4.16, 95% CI 3.41–5.09, p<0.001). This association was significantly modified by arrest location (p<0.001 for interaction). In residential settings, AEDs were applied in 11.0% (136/1,240) of cases where a volunteer responder arrived, compared with 1.2% (176/15,124) of cases without a volunteer responder (aOR 7.38, 95% CI 5.81–9.38). In non-residential settings, AED application occurred in 38.0% (60/158) of cases with a volunteer responder versus 19.4% (953/4,917) without (aOR 1.71, 95% CI 1.22–2.40).

In addition, the effect of age on AED application was also modified by arrest location (p=0.01 for interaction), with adjusted odds ratios showing that older age was associated with lower likelihood of AED use in residential settings, whereas age had little impact in non-residential settings (Figure 5).

**Figure 5.**
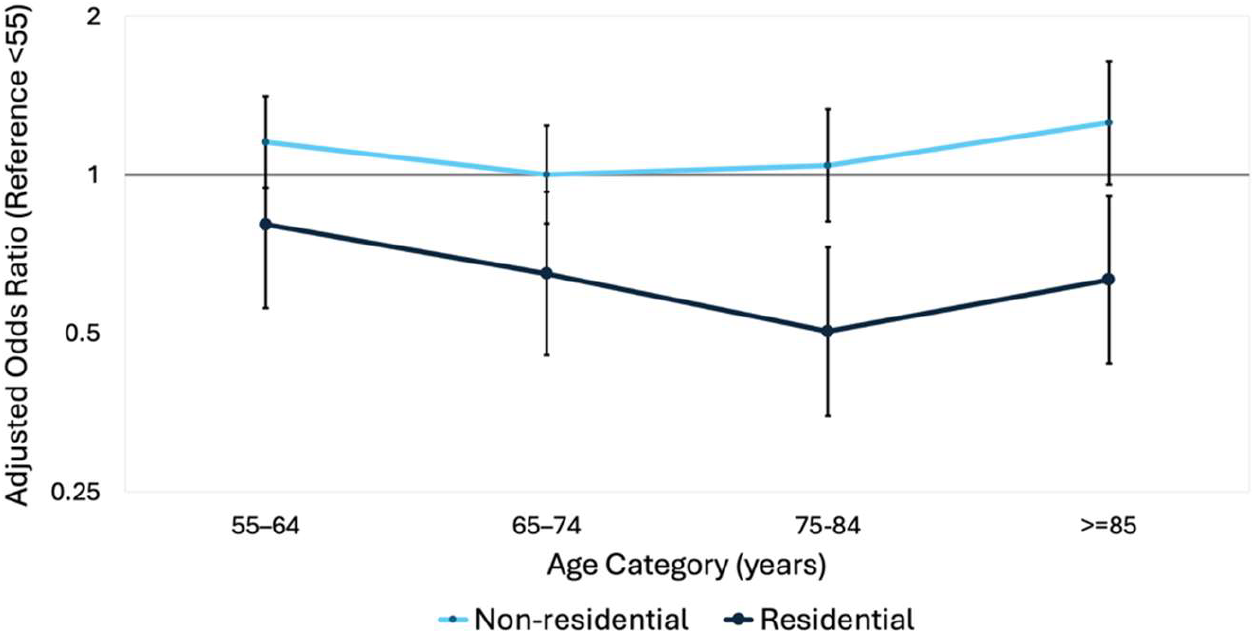
Arrest location modifies the effect of age on BAED application. Adjusted odds ratios (ORs) with 95% confidence intervals (CIs) are shown for different age categories (<55 years as reference). The light blue line represents non-residential arrests, and the dark blue line represents residential arrests. Error bars indicate 95% CIs. The effect of age on AED application is modified by arrest location, with residential settings showing generally lower odds across older age groups.

### Clinical Outcomes

In outcome analyses, both bystander cardiopulmonary resuscitation (CPR) and BAED application were strongly associated with improved clinical outcomes (Table 2). Bystander CPR was associated with higher odds of favorable neurological outcome (aOR 2.17, 95% CI 1.73– 2.73) and survival (aOR 1.62, 95% CI 1.37–1.92), as well as increased likelihood of prehospital ROSC (aOR 1.41, 95% CI 1.26–1.57).

**Table 2.**
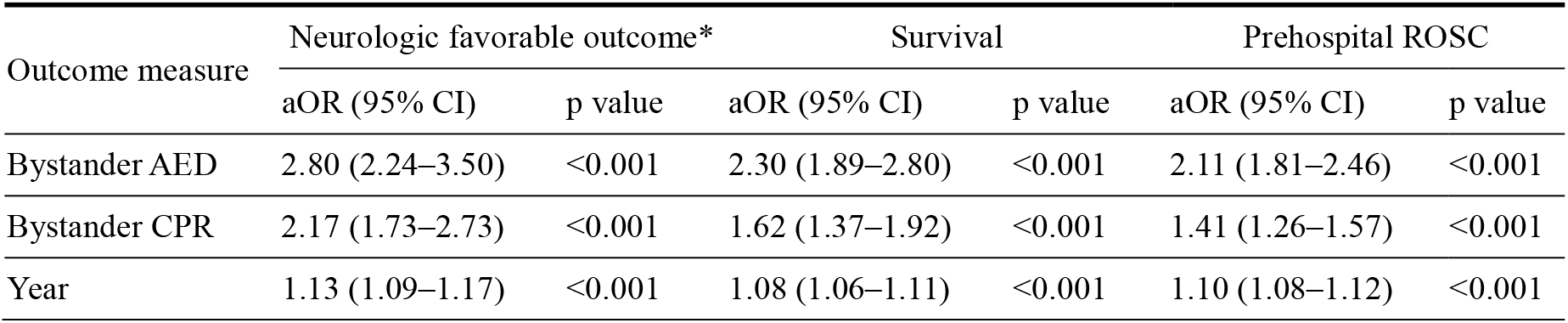
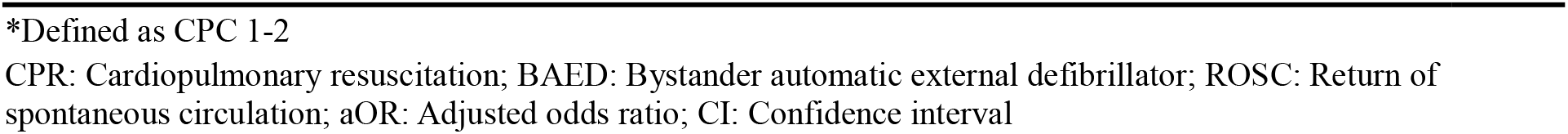
Association between bystander interventions, calendar year, and clinical outcomes. Odds ratios from logistic regression analyses examining the association of bystander cardiopulmonary resuscitation (CPR), bystander automatic external defibrillator (BAED) application, and year with favorable neurologic outcome (CPC 1–2), overall survival, and prehospital return of spontaneous circulation (ROSC) among adult (age ≥ 18) out-of-hospital cardiac arrest cases. Covariates include gender, age, ethnicity, weekend, time of day, location of arrest, witness status, and emergency medical services response time.

| Outcome measure | Neurologic favorable outcome* |  | Survival |  | Prehospital ROSC |  |
| --- | --- | --- | --- | --- | --- | --- |
|  | aOR (95% CI) | p value | aOR (95% CI) | p value | aOR (95% CI) | p value |
| Bystander AED | 2.80 (2.24–3.50) | <0.001 | 2.30 (1.89–2.80) | <0.001 | 2.11 (1.81–2.46) | <0.001 |
| Bystander CPR | 2.17 (1.73–2.73) | <0.001 | 1.62 (1.37–1.92) | <0.001 | 1.41 (1.26–1.57) | <0.001 |
| Year | 1.13 (1.09–1.17) | <0.001 | 1.08 (1.06–1.11) | <0.001 | 1.10 (1.08–1.12) | <0.001 |

\*Defined as CPC 1-2
CPR: Cardiopulmonary resuscitation; BAED: Bystander automatic external defibrillator; ROSC: Return of spontaneous circulation; aOR: Adjusted odds ratio; CI: Confidence interval

BAED application demonstrated an even stronger association with outcomes, including favorable neurological outcome (aOR 2.80, 95% CI 2.24–3.50), survival (aOR 2.30, 95% CI 1.89–2.80), and prehospital ROSC (aOR 2.11, 95% CI 1.81–2.46). These associations corresponded to a relatively low NNT; only 8.7 (95% CI 7.5–10.4) BAED applications were required to achieve one additional favorable neurological survival. The NNT for survival to discharge and prehospital ROSC were 8.2 (95% CI 7.0–9.7) and 5.7 (95% CI 5.0–6.6), respectively.

The clinical benefit of BAED application remained consistent across multiple subgroups, with no significant effect modification by arrest location (p=0.41), witness status (p=0.42), or myResponder volunteer arrival (p=0.31). We observed a similar pattern regarding the time of day, with a significant survival benefit during the day (aOR 3.26, 95% CI 2.54–4.18) and evening (aOR 2.84, 95% CI 1.92–4.19). While the point estimate for nighttime applications was neutral (aOR 1.00, 95% CI 0.39–2.58), the wide confidence interval, driven by the low volume of AED applications during these hours, suggests this result is underpowered.

Across the study period, temporal improvements in outcomes were observed, with increasing odds of favorable neurological outcome (aOR per year 1.13, 95% CI 1.09–1.17), survival (aOR per year 1.08, 95% CI 1.06–1.11), and prehospital ROSC (aOR per year 1.10, 95% CI 1.08–1.12).

## Discussion

In this large, population-based ‘living laboratory’ study of over 20,000 out-of-hospital cardiac arrests in Singapore, we observed a sustained and substantial increase in BAED application over an 11-year period from 1.7% to 9.6% of OHCA cases. This improvement paralleled the implementation of multiple coordinated public health interventions, including widespread AED deployment in both public and residential locations, national training initiatives, and the introduction of smartphone-activated volunteer responders.^12–16^ The progressive increase in BAED applications over time, independent of other covariates, suggests that Singapore’s multi-pronged strategy to increase BAED has been effective. These findings align with prior studies showing that community-based interventions can improve bystander action in OHCA.^2,8,9,17,18^

Increasing BAED application was strongly associated with improved clinical outcomes, including nearly a threefold increase in favorable neurological survival and a NNT of 8.7, consistent with previously reported values.^4^ Importantly, the survival advantage from AED application was stable across arrest settings and patient subgroups, with no evidence of effect modification by location, witness status, or volunteer responder involvement. This indicates that while the likelihood of BAED application may vary by context, its clinical effectiveness remains robust. It must also be acknowledged that the proportion of patients with an initial shockable rhythm was substantially higher in the BAED group. However, this difference is likely related to the fact that patients who receive bystander AED application undergo earlier rhythm assessment, increasing the likelihood that a shockable rhythm is identified before degeneration to non-shockable states. In this context, initial rhythm is not an independent baseline characteristic but is partially determined by the timing of rhythm assessment, which itself lies on the causal pathway between BAED application and improved outcomes. As such, the higher prevalence of shockable rhythms in the BAED group would be an expected mediator of treatment effect rather than a source of bias.^4^

Smartphone-activated volunteer responder arrival was strongly associated with increased BAED application (aOR 4.16, p<0.001), with an even greater effect in residential settings (aOR 7.38). Although most arrests occur in residential locations (76% in this cohort), increasing BAED in these settings has been a persistent challenge globally in OHCA care.^22,23,30–32^ However, over the study period, the residential BAED rate in Singapore increased from 0.1% to 4.6%, suggesting meaningful progress. These findings indicate that pairing residential AED placement with smartphone-activated volunteer responder systems may help overcome longstanding barriers and extend the reach of early defibrillation into the home.^32,33^

While the overall trajectory of BAED applications in Singapore is encouraging, our results also identify opportunities to further optimize bystander response. BAED application remained less likely among female patients, which is consistent with prior reports of sex-based differences in BAED application observed in other regions.^19,21,34,35^ Our findings extend this evidence to a Southeast Asian context and suggest that sex-based differences in BAED application are not system-specific but may reflect broader, transnational barriers to bystander intervention.^36,37^ The literature on bystander CPR hesitation indicate that barriers include concern over inappropriate physical contact, fear of social or legal consequences, and apprehension about causing harm.^38^ Qualitative and simulation-based research further suggests that hesitation to remove clothing from female patients, shaped by prevailing social norms, may contribute to sex-based differences.^39,40^ Existing social barriers are exacerbated by the lack of female representation in AED training materials and equipment.^40,41^ Collectively, these findings highlight the global need for resuscitation training and public education that explicitly address social norms, fears, and misconceptions that may disproportionately deter BAED use in women.

In residential settings, the odds of BAED application declined progressively with increasing age relative to patients aged <55, reaching a nadir in the 75–84-year group before a modest rise among those aged ≥85. In contrast, in non-residential settings, BAED application remained largely stable across age strata, with adjusted odds close to the <55 reference group. This divergence implies that age-related reductions in BAED application are concentrated in residential arrests, consistent with setting-specific barriers, such as fewer available bystanders and access constraints. This population may benefit from targeted interventions such as the intentional recruitment of myResponder volunteers from communities with a higher median age.

Delayed EMS response and Malay ethnicity were associated with increased BAED application. Longer EMS response may provide more time and opportunity for bystanders to retrieve and apply an AED.^21^ Higher BAED application in Malay individuals may be reflecting socio-demographic factors such as higher residence in public housing (96%), where residential AED deployment is concentrated, and larger household sizes that facilitate CPR and AED retrieval.^42^ Despite this, Malay patients have poorer OHCA outcomes than members of other ethnic groups in comparable arrest scenarios, suggesting that other factors may attenuate the survival benefit of increased AED use.^43^

The findings of this study have several important implications for policy and practice. Efforts to improve OHCA outcomes should consider targeting residential BAED rates as most arrests occur in residential locations, BAED application is just as strongly associated with improved neurological in residential locations as in public, and residential locations are a current gap in care with disproportionately lower rates of BAED application. A combined approach of expanded AED availability in high-density housing, alongside smartphone-activated volunteer responder system integration, can serve as a model for improving residential BAED rates.^44^ Additional opportunities to further BAED strategies include increasing female representation in AED training materials and conducting mixed-methods research to understand the barriers to BAED application in females.

A few limitations should be considered. Volunteer responder activation was recorded through user interaction with the myResponder application, and missing data likely led to under-ascertainment, with some cases classified as having no responder despite their presence; this would bias results toward underestimating the benefit of volunteer responders. In addition, as myResponder was only implemented from 2016 onward, earlier cases were necessarily classified as having no volunteer response, meaning comparisons reflect periods with and without the system in place, which may overestimate the effect. Additional confounding from unmeasured factors such as AED proximity, responder density, or bystander training may also influence results, potentially biasing estimates in either direction depending on their distribution. Exclusion of cases with missing data may introduce selection bias, although the relatively small proportion suggests limited impact. Finally, these findings may not be fully generalizable to regions with differing emergency response systems or different population structures. However, given the well-established benefit of bystander AED use across diverse settings, differences are more likely to relate to the magnitude of effect and implementation context rather than the direction of benefit.

## Conclusion

Our study demonstrates that sustained population-level efforts improved BAED application over the years in Singapore. This improvement highlights the effectiveness of an integrated public health approach combining widespread AED deployment and national training with smartphone-activated volunteer responders. In residential arrests, volunteer responder arrival strongly improved the BAED application likelihood, indicating an important pathway for extending early defibrillation into the home. Overall, Singapore’s model offers a scalable blueprint for other urban settings, while further gains may require targeted, community-integrated interventions to close remaining gaps in BAED application.

## Data Availability

The data analyzed during the study are available from the corresponding author (via email) upon reasonable request.

## Acknowledgements

The author would also like to thank Ms Maeve Pek from Pre-hospital and Emergency Research Centre, Duke-NUS Medical School, Singapore; the late Ms Susan Yap from Department of Emergency Medicine, Singapore General Hospital, Singapore; Ms Noor Azuin, Ms Nurul Asyikin and Ms Liew Le Xuan from Unit for Pre-hospital Emergency Care, Singapore; Ms Charlene Ong and Ms Anju Devi from Accident & Emergency, Changi General Hospital, Singapore and Ms Woo Kai Lee from Department of Cardiology, National University Heart Centre Singapore for their contributions and support to the Singapore PAROS registry, and Unit for Pre-hospital Emergency Care, Singapore for facilitating/implementing the initiatives that have brought about improved response to OHCA.

## Singapore PAROS Investigators

Desmond R Mao, Khoo Teck Puat Hospital, Singapore; Han Nee Gan, Changi General Hospital, Singapore; Wei Ming Ng, Ng Teng Fong General Hospital, Singapore; Nausheen D Edwin, Sengkang General Hospital, Singapore; Shir Lynn Lim, National University Heart Centre Singapore, Singapore; Ling Tiah, Changi General Hospital, Singapore, Singapore; Wei Ling Tay, Ng Teng Fong General Hospital, Singapore; Yih Yng Ng, National University of Singapore, Singapore; Si Oon Cheah, Urgent Care Clinic International, Singapore; Shun Yee Low, Sengkang General Hospital, Singapore; E Shaun Goh, Woodlands Hospital, Singapore; Gayathri D Nadarajan, Singapore General Hospital, Singapore; Ivan SY Chua, Singapore General Hospital, Singapore; Andrew FW Ho, Singapore General Hospital, Singapore.

## Disclosures

Prof Marcus EH Ong reports grants from the Laerdal Foundation, Laerdal Medical, and Ramsey Social Justice Foundation for funding of the Pan-Asian Resuscitation Outcomes Study; an advisory relationship with Global Healthcare SG, a commercial entity that manufactures cooling devices. He has a licensing agreement with ZOLL Medical Corporation and patent filed (Application no: 13/047,348) for a “Method of predicting acute cardiopulmonary events and survivability of a patient.” He is also the co-founder and scientific advisor of TIIM Healthcare, a commercial entity which develops real-time prediction and risk stratification solutions for triage. These organizations have no role in conducting this research.

## Declaration of Interest

Prof Marcus EH Ong is a member of the Editorial Board of Resuscitation.

## Funding Sources

This study was funded by National Medical Research Council, Singapore, through the Clinician Scientist Awards (NMRC/CSA/024/2010, NMRC/CSA/0049/2013 and NMRC/CSA-SI/0014/2017) and by the Ministry of Health, Singapore, through the Health Services Research Grant (HSRG/0021/2012), awarded to Prof Ong.

## Role of the Funding Source

The funders of the study had no role in the study design, data collection, data analysis, data interpretation, or writing of the report.

